# A Mixed-Methods Feasibility Study of a Novel AI-Enabled, Web-Based, Clinical Decision Support System for the Treatment of Major Depression in Adults

**DOI:** 10.1101/2022.01.14.22269265

**Authors:** Sabrina Qassim, Grace Golden, Dominique Slowey, Mary Sarfas, Kate Whitmore, Tamara Perez, Elizabeth Strong, Eryn Lundrigan, Marie-Jeanne Fradette, Jacob Baxter, Bennet Desormeau, Myriam Tanguay-Sela, Christina Popescu, Sonia Israel, Kelly Perlman, Caitrin Armstrong, Robert Fratila, Joseph Mehltretter, Karl Looper, Warren Steiner, Soham Rej, Jordan F. Karp, Katherine Heller, Sagar V. Parikh, Rebecca McGuire-Snieckus, Manuela Ferrari, Howard Margolese, David Benrimoh

## Abstract

The objective of this paper is to discuss perceived clinical utility and impact on physician-patient relationship of a novel, artificial-intelligence (AI) enabled clinical decision support system (CDSS) for use in the treatment of adults with major depression. Patients had a baseline appointment, followed by a minimum of two appointments with the CDSS. For both physicians and patients, study exit questionnaires and interviews were conducted to assess perceived clinical utility, impact on patient-physician relationship, and understanding and trust in the CDSS. 17 patients consented to participate in the study, of which 14 completed. 86% of physicians (6/7) felt the information provided by the CDSS provided a more comprehensive understanding of patient situations and 71% (5/7) felt the information was helpful. 86% of physicians (6/7) reported the AI/predictive model was useful when making treatment decisions. 62% of patients (8/13) reported improvement in their care as a result of the tool. 46% of patients (6/13) felt the app significantly or somewhat improved their relationship with their physicians; 54% felt it did not change. 71% of physicians (5/7) and 62% of patients (8/13) rated they trusted the tool. Qualitative results are analyzed and presented. Findings suggest physicians perceived the tool as useful in conducting appointments and used it while making treatment decisions. Physicians and patients generally found the tool trustworthy, and it may have positive effects on physician-patient relationships.

## Introduction

Clinical Decision Support Systems (CDSS; Sim et al., 2001) in mental health treatment, including systems that incorporate artificial intelligence (AI) to assist with treatment management, are being developed and researched at an increasing rate (Tran et al., 2019). CDSS are tools that serve to assist clinicians in making accurate and informed decisions regarding treatment and are meant to supplement the clinician-patient relationship (Patel et al., 2008). In this paper we discuss Aifred, a novel CDSS that aims to support clinicians and patients as they make choices about the treatment of major depression.

The Aifred CDSS is described in detail in previous work (Benrimoh et al., 2020; Popescu et al., 2021). Briefly, it is a web-based platform accessible from mobile or desktop devices and provides both clinicians and patients with an access portal. Patients use the system to respond to standardized questionnaires and track their questionnaire scores and treatments over time. Physicians have access to the same questionnaire and treatment data, assisting them in the practice of measurement-based care (MBC; Goldberg et al., 2018; Scott & Lewis, 2015) Additionally, physicians have access to a clinical treatment algorithm, an operationalized version of the 2016 CANMAT guidelines for the treatment of major depressive disorder (Kennedy et al. 2016). This rule-based system considers where patients are in treatment and situates them within the guidelines, assisting clinicians in the use of algorithm-guided treatment (AGT; Adli et al., 2017; Bauer et al., 2009; Yoshino et al., 2009). Finally, the Aifred CDSS incorporates a novel element: an artificial intelligence (AI)-based predictive model. This model inputs patient clinical and demographic characteristics to generate treatment-specific remission probabilities for a number of commonly prescribed antidepressants. Pharmaceutical treatments are presented to the physician alongside the remission probability for each medication for which the model has data and other guideline-derived clinical information, such as dosing or specific symptoms for which the treatment may have evidence of effectiveness (e.g., fatigue, sleep or pain) (Kennedy et al., 2016). The CDSS is designed to be used during sessions with patients and to assist in shared decision-making (Benrimoh et al., 2020). The rationale for including the AI component is that personalization of treatment in depression remains a challenge, often relying on trial and error. As such, a point-of-care predictive model may enable identifying optimally effective treatments for patients earlier in their care (Benrimoh et al., 2018).

The Aifred CDSS, or simply “tool”, has previously been shown to be easy to use by clinicians and perceived as clinically useful and trustworthy in a simulation center study (Benrimoh et al., 2020; Tanguay-Sela et al., 2021). The current paper reports on the results of an in-clinic Feasibility study of the Aifred CDSS. The primary outcome of this study was clinical feasibility, as demonstrated by no increase in appointment length (Popescu et al., 2021). This paper considers both quantitative and previously unreported qualitative results from this study, with a focus on discussing perceived clinical utility, trust, and impacts on the clinician-patient relationship of the Aifred CDSS.

## Methods

Ethics approval for the study was obtained by the Research Ethics Boards of the Douglas Mental Health University Institute (identifier: NCT04061642). The study was carried out according to the Tri-Council Policy Statement on the Ethical Conduct for Research Involving Humans (Tri-Council Policy Statement, 2005), with participants providing written informed consent.

The study was a single arm, naturalistic follow-up study aimed at assessing the acceptability and useability of the software, conducted from January to November 2020. The study sample consisted of two populations: 1) physicians, including psychiatrists and family physicians and 2) patients of these physicians. Patients were eligible if they were over the age of 18, capable of providing informed consent, diagnosed by their physician with an active major depression using DSM-5 criteria (APA, 2013) which the clinician felt was the primary mental disorder being treated, and did not have bipolar disorder. 17 patients consented to participate in the study, of which 14 completed, and qualitative data was obtained for 12 participants. Detailed methods can be found in Popescu et al., 2021.

The study was conducted at university hospitals, primary care, and psychiatric clinics in Québec, Canada. Participating physicians were invited to an introductory session to learn how the AI model was created and trained on how to use the CDSS. Patients who met the eligibility criteria and provided informed consent had accounts created that were linked to their respective physicians. Due to COVID-19 and public health recommendations, the study was adapted to be completed entirely via telemedicine early in the course of the study.

After patients provided informed consent, their next appointment with their physician served as the baseline appointment. It is important to note that the tool was not used by the physician during this appointment. This was because the primary outcome for the study was change in appointment length, and this design allowed the measurement of appointment length pre- and post-CDSS introduction. Patients were then seen at least two subsequent appointments, termed visit 1 and visit 2, during which the tool was used by the physician. At baseline, patients completed a number of questionnaires including a demographics questionnaire, a Patient Health Questionnaire-9 (PHQ-9), a General Anxiety Disorder-7 (GAD-7), an Alcohol Use Disorders Identification Test (AUDIT), a Drug Abuse Screen Test (DAST), and a Standardized Assessment of Personality-Abbreviated Scale Self-Assessment (SAPAS) (Kroenke et al., 2001; Spitzer et al., 2006; Bohn et al., 1995; Skinner, 1982; Merlhiot, 2014). Subsequently, on a weekly basis patients were asked to complete the PHQ-9 and GAD-7 to assess depressive and anxious symptoms, as well as the Patient-Rated Inventory of Side Effects-20 (PRISE-20) and Frequency, Intensity, and Burden of Side Effects Rating (FIBSER), which, together, allow for the assessment of the presence, frequency, intensity, and burden of medication side effects (Katz et al., 2012; Wisniewski et al., 2006). Patients were reminded to complete questionnaires by automated email notifications.

Following visit 2, patients were administered the Scale to Assess Therapeutic Relationships in Community Mental Health Care - Patient Version (STAR-P) to study the quality of the patient-physician relationship as perceived by each patient (McGuire-Snieckus et al. 2007), as well as a customized exit questionnaire and custom semi-structured interview to explore the patient experience with the tool using a quantitative and qualitative approach, respectively. After all of a physician’s patients had completed visit 2, they were similarly administered the Scale to Assess Therapeutic Relationships in Community Mental Health Care - Clinician Version (STAR-C) (McGuire-Snieckus et al. 2007), as well as a customized exit questionnaire and custom semi-structured interview, all of which explored concepts akin to the patient versions but from a clinician’s perspective. During all interviews, research assistants (RAs) took extensive written notes and these formed the dataset for the qualitative analysis. Clinicians and patients were compensated for their time. The study flow was documented in Popescu et al., 2021 (Figure 1).

## Analysis

### Quantitative

Descriptive statistics are presented for the demographics questionnaire, baseline clinical questionnaires, STAR and customized exit questionnaire. Quantitative results regarding main study outcome (appointment length) can be found in Popescu et al., 2021.

### Qualitative Analysis

Written notes obtained during the 20-30 minute patient and physician semi-structured interviews were reviewed to extract initial themes prior to data analysis, as a means of commencing the codification of data while minimizing bias. RAs met patients and clinicians one-on-one and asked a set of questions, with instructions to record participant responses in writing; follow-up questions were permitted for clarification or to more fully explore a participant’s comment. RAs were familiarized with the data and extracted an initial set of categories and themes based on the experiences of patients and physicians in the study. RAs then reviewed the data once more to refine the initial themes extracted, which consisted of collapsing, splitting, eliminating and creating new themes. Five categories were identified: 1) physician; 2) patient; 3) physician-patient relationship; 4) COVID-19 and 5) miscellaneous. Themes were identified within the categories. Overarching themes encompassed similar concepts, spanning multiple participants, as found within the data. Themes could be further separated into sub-themes. The physician category consisted of 16 themes, the patient category had 15 themes, both the physician-patient relationship and COVID-19 categories had 5 themes and the miscellaneous category had 2 sub-themes (see Supplementary Material for sub-themes). Using an inductive thematic analysis approach and following the steps laid out in Braun & Clark, 2006, data was coded without trying to fit it into a pre-existing coding frame or the researcher’s analytic preconceptions, and allowed the themes identified to be strongly linked to the data themselves.

RAs independently read and coded excerpts of the data into subheadings of the thematic table. All RAs were assigned the data corresponding to all participants, so that each participant’s data was independently coded by all RAs. This data was then condensed by collapsing some of the themes and rearranging the subheadings to reduce the overlap of excerpts between subheadings. The RAs then independently reread and coded all of the data into a final summary table. This stage ensured that any data that had been missed in earlier coding stages could be added, and also validated the new themes in relation to the full data set. Finally, data were arranged into four final categories of interest to the central themes of this paper (clinical utility and clinician-patient relationship). These were Clinical Utility and Useability of the App, Impact on Physician-Patient Interaction and Relationship, Experience of Measurement-Based Care, and Understanding and Trust.

The main qualitative research question was: how do patients and clinicians perceive the clinical utility and trust in the tool, and its impact on the clinician-patient relationship?

## Results

### Quantitative Results

#### Sample Description

Ten physicians consented to participate in the study; however 3 were unable to recruit patients due to interruptions related to COVID-19 in their clinical practice. As such, the final physician sample consisted of 7 physicians (4 general practitioners, 3 psychiatrists). Twenty patients were approached to join the study, with 17 consenting to participate. One patient dropped out before the baseline appointment and 2 dropped out after the baseline appointment but before visit 1 and therefore did not interact with the CDSS and their physician together in a session. This resulted in 14 patients (9 women and 5 men) completing the study (defined as completing visit 2). Mean participant age was 36.43 years (SD 14.84). See table 1 for patient demographics and table 2 for baseline clinical questionnaire scores.

**Table 1.** Patient demographic characteristics.

| Patient Characteristics | n (%) | Mean (SD) |
| --- | --- | --- |
| **Age** (years; n=14) | 14 (N/A) | 36.43 (14.84) |
| **Sex** (n=14)<br>Male<br>Female | 5(35.71)<br>9(64.29) |  |
| **Gender** (n=14)<br>Men<br>Women | 5(35.71)<br>9(64.29) |  |
| **Ethnicity** (n=13)<br>Caucasian<br>Caribbean<br>African or African American<br>Unanswered | 10(76.92)<br>1(7.69)<br>1(7.69)<br>1(7.69) |  |
| **Residency Status** (n=13)<br>Canadian citizen<br>Immigrant status (>5 years ago)<br>Immigrant status (<5 years ago) | 11(84.62)<br>1(7.69)<br>1(7.69) |  |
| **Adoption Status** (n=13) |  |  |
| Not adopted | 12(92.31) |  |
| Adopted | 1(7.69) |  |
| **Employment Status** (n=13) |  |  |
| Full-Time | 7(53.85) |  |
| Part-Time | 1(7.69) |  |
| Disability (not working) | 2(15.38) |  |
| Unemployed + volunteer work | 2(15.38) |  |
| Unemployed | 1(7.69) |  |
| **Relationship Status** (n=13) |  |  |
| Married | 4(30.77) |  |
| Divorced | 1(7.69) |  |
| Dating a single partner | 2(15.38) |  |
| Not in a relationship | 6(46.15) |  |
| **Highest Level of Education**<br>n=12) |  |  |
| Master's degree | 3(25.00) |  |
| Bachelor's degree | 1(8.33) |  |
| University, no degree | 3(25.00) |  |
| Collège d'enseignement général<br>et professionnel | 3(25.00) |  |
| High school or equivalent (GED) | 2(16.67) |  |
| **Years of Education** (n=12) | 12(N/A) | 15 (6.42) |
| **Income** (\$ CA) (n=10) | 10(N/A) | 82333.33<br>70099.93) |

**Table 2.** Baseline questionnaire scores.

|  |  |  |
| --- | --- | --- |
| Questionnaire Baseline Score<sup>1</sup> | Frequency,<br>n (%) | Mean (SD) |
| **GAD-7** (n=14)<br>0-4: no or minimal anxiety<br>5-9: mild anxiety<br>10-14: moderate anxiety<br>15-21: severe anxiety | 1(7.14)<br>4(28.57)<br>3(21.43)<br>6(42.86) | 12.21 (5.81) |
| **PHQ-9** (n=15)<br>0-4: minimal or no depression<br>5-9: mild depression<br>10-14: moderate depression<br>15-19: moderately severe depression<br>20-27: severe depression | 1(6.67)<br>1(6.67)<br>4(26.67)<br>7(46.67)<br>2(13.33) | 14.80 (5.61) |
| **SAPAS-SA** (n=15)<br>< 3 points: negative screen for personality disorder<br>≥ 3 points: positive screen for personality disorder. Further clinical evaluation is warranted. | 5(33.33)<br>10(66.67) | 3.53 (2.23) |
| **DAST-10** (n=15)<br>0: no problems reported<br>1-2: low level<br>3-5: moderate level<br>6-8: substantial level<br>9-10: severe level | 8(53.33%)<br>3(20.00)<br>0(0.00)<br>3(20.00)<br>1(6.67) | 2.40 (3.60) |
| **AUDIT** (n=15)<br>< 8: negative screen for harmful alcohol use<br>≥ 8: positive screen for harmful alcohol use | 12(80.00)<br>3(20.00) | 4.40 (3.58) |
| **QIDS** (n=12)<br>1-5: no depression<br>6-10: mild depression<br>11-15: moderate depression<br>16-20: severe depression<br>21-27: very severe depression | 2(16.67)<br>2(16.67)<br>3(25.00)<br>4(33.33)<br>1(8.33) | 13.12 (6.04) |
| **WHODAS** (n=13) | N/A | 54.62 (27.08) |
| **ACE** (n=13)<br>0 adverse childhood events<br>1 adverse childhood event<br>2 adverse childhood events<br>3 adverse childhood events<br>4+ adverse childhood events | 4(30.77)<br>3(23.08)<br>1(7.69)<br>4(30.77)<br>1(7.69) | 1.62 (1.45) |
| **IDS-C** (n=13)<br>0-11: no depression<br>12-23: mild depression<br>24-36: moderate depression<br>37-46: severe depression<br>47-84: very severe depression | 0(0.00)<br>4(30.77)<br>5(38.46)<br>3(23.08)<br>1(7.69) | 30.00 (12.88) |

### Descriptive Results

#### Physician-Patient Relationship

In order to determine the quality of the patient-physician relationship, physicians (n=7) completed the STAR-C and patients (n=13) completed the STAR-P at the end of the study. The questionnaire was not administered at the beginning of the study given the range of patient-physician relationship qualities and duration that could not be controlled for due to the small sample size. At the end of the study, patients had a mean total score of 33.62 (SD 2.90) and physicians had a mean total score of 31.14 (SD 2.63). In the original STAR study, patients had a mean total score 38.4 (SD 12.0) and physicians had a mean total score of 31.5 (SD 6.9). The maximum score for both questionnaires is 48, with higher scores indicating a stronger relationship. The mean STAR-P subscores for positive clinical input, positive collaboration and non-supportive clinical input were 10.23 ± 1.88, 21.92 ± 2.24 and 10.54 ± 2.58, respectively, compared to 9.3 ± 3.0, 19.9 ± 6.7 and 9.3 ± 3.3, the subscores in the original STAR-P. The mean STAR-C subscores for positive clinician input, positive collaboration and emotional difficulties were 9.93 ± 1.38, 17 ± 2.60 and 10.57 ± 1.79, respectively, compared to 8.9 ± 1.6, 7.4 ± 2.7 and 15.3 ± 4.0, the subscores in the original STAR-C. As such, after using the tool there were no marked differences in our sample with respect to clinician-patient relationship compared to a reference population. In addition, the customized exit questionnaire was completed by 13 patients. 46% of the patients (6/13) felt that the app significantly or somewhat improved the patient-physician relationship and the other 7 (54%) felt that it did not change.

#### Clinical Utility

86% of the physicians (6/7) reported that the information provided by the CDSS was useful for getting a better sense of their patients’ situations and 71% (5/7) felt that the information provided by the tool was helpful, indicated by a 4 or 5 on the 5-Point Likert scale. 71% (5/7) reported that they believed they would continue to use this application after the study and 29% (2/7) reported that they would not. 57% of physicians (4/7) reported that they would use the AI/predictive model in future clinical practice, and the other 43% (3/7) was unsure.

Importantly, 86% of the physicians (6/7) thought the AI/predictive model was somewhat or very useful in helping them make their treatment decisions, indicated as a 4 or 5 on the 5-Point Likert scale, and one physician was unsure about its usefulness. 71% (5/7) of the physicians rated that they trusted the tool, indicated as a 4 or 5 on the 5-Point Likert scale.

As indicated by scores of 4 and 5 on the 5 point Likert scale, 62% of the patients (8/13) rated that they trusted the tool. In addition, 62% of patients (8/13) felt that the tool allowed them to fully report how they were feeling and 62% of patients (8/13) reported that the questions covered the things most important to them. Furthermore, 77% (10/13) of patients gave scores of 4 and 5 in response to whether keeping track of their symptoms and treatments in the tool helped their physician better manage their care. Overall, 62% of patients (8/13) felt the app improved their care and the other 38% (5/13) thought the tool had no effect. No patients noted that their care worsened as a result of the tool.

### Qualitative Results

All physicians (7/7) who recruited patients in the study completed a final interview. In addition, 12 of the 14 patients who completed the study completed an end interview; of these, 7 patients were patients of GPs and 5 were patients of psychiatrists. 2/14 patients could not be contacted to complete a final interview. Below are the data from these interviews, organized into four categories: 1) Clinical utility and useability of the application; 2) Physician-patient interaction and relationship; 3) Experience of measurement-based care; and 4) Understanding and trust. Sub-categories are presented in each category. Note that when a certain number of participants are noted as making a statement, this does not imply that the remainder of participants made the opposite statement.

#### A. Clinical Utility and Useability of the App

##### Ease of Use

Overall, 10 of 12 patients and 6 of 7 physicians reported that the app was easy to use. Despite noting the tool was easy to use, 3 physicians reported they would have felt more comfortable with the tool with more time and practice. Further comments regarding ease of use can be found in the supplementary material and Box 1.

###### Box 1.

Physician and Patient Quotes:

*Clinical Utility and Useability of the App*

Physicians and patients shared comments regarding ease of using the app, as well as its clinical utility.

Physician 1 reported the tool was “easy and user-friendly, steps and suggestions are well explained.”

Physician 2 shared that “reading the change in scores [prior to appointments] definitely gave some orientation as to where the patient is going and if things are improving or not”

Patient 1 felt the tool “allowed [the] doctor to understand symptoms more when she couldn’t verbalize how she was feeling.”

Patient 2 also noted that they would continue to use the tool in a maintenance capacity, sharing they “would use it but a couple of times per year… [to track] mood changes, [but] not week to week”

*Impact on Physician-Patient Interaction and Relationship*

Physicians and patients agree that the tool improved patient-clinician interactions.

Patient 3 reported feeling the tool “would be very useful for a new patient-physician relationship or a doctor without much time to give them info on when they’re not doing well and to take them seriously.”

Physician 3 stated they “felt like there was another outside kind of help and of objectification and that helpful the rapport with the patient” The tool also improved patient engagement in their own care.

Multiple patients reported they were “better able to self-reflect and articulate how [they were] feeling at visits”

*Measurement-Based Care: Patient Perspective*

Patients reported the tool brought more awareness to their changing thoughts and feelings.

Patient 2 shared they were able to “reflect on how [their] past week has been [and] helped her remember her moods.”

Patient 3 stated “mood really fluctuates during depression so I think it really helped to point out that there were days where I felt ok,”

Patient 4 shared “there’s actually something nice about filling in these questionnaires more often. The questions always feel a little off the first time so doing it multiple times takes away the stress of having something riding on me to do it right. […] [I] like that this is a neutral way to be asked about your feelings without having a person or putting on a front for others (such as friends). I enjoyed it, filling in the questionnaires was a useful therapy.”

Patient 5 reported: “I know how I feel. I know. And I’m fed up with it; It makes me realize certain things that I was ignoring. When you’re like this, you don’t think about what you feel. You know you’re different, but this helps you realize how you are; It helped me to realize what it is from the questions, and make me realize what I have. I don’t know about depression. You hear about people with depression, but you don’t know anything about it until you have it. Even the people at work knew because they saw I was not the same, but I did not realize it.”

*Understanding and Trust*

Physicians generally reported trusting the tool enough to integrate it into their care.

Physician 4 shared the tool suggestions “were reasonable and mirrored guidelines”

Patients shared little concern regarding their understanding of the tool.

Patient 4 reported “As long as it works, it works – whether it’s magic or AI”

##### Clinical Utility

With respect to clinical utility as perceived by patients, some patients were able to identify how their questionnaire responses were being used by their physician, with one patient expressing that if they “reported higher anxiety levels (for example) that week, [her] doctor saw that in [the] appointment when [they] looked at tool so [they] increase[d] medication.” 3 of 12 patients (2 patients of GPs; 1 patient of a psychiatrist) commented that the tool did not necessarily have an effect on their care as they had already been on medication. This was expected as part of the study design, where the tool was introduced after the baseline visit (and therefore after pharmacotherapy was likely to have been initiated).

Patients reported how they used questionnaire responses and seeing their trends graphed to facilitate better communication between themselves and their care provider. One patient shared that it helped in “highlighting my priorities”. Another patient (of a GP) stated that “I think what it accomplished was to give me the info I needed to transmit to my GP when I spoke to him. I could look back and reflect on how I felt. That info being there in black and white for me made it easier to be prepared to speak to him.” Another patient (of a GP) discussed how the tool “allowed [the] doctor to understand symptoms more when she couldn’t verbalize how she was feeling.”

Physicians commented on the clinical utility of the CDSS, with particular emphasis on how they interpreted and used the tool’s clinical algorithm. One doctor described that it was “nice to objectify scores, as [they] were able to explain what [they] were doing, [and] told the patient what the algorithm suggested”.Another physician described their treatment decisions using the tool in the context of the CANMAT guidelines, saying that it is “helpful to know that clinical decisions were supported by CANMAT guidelines, not just physician recommendations and/or opinions”. Interestingly, 4/7 (2 Psychiatrists, 2 GPs) doctors shared comments regarding their openness to prescribe medications they may not otherwise have considered based on the information presented within the algorithm module.

A trend that emerged from physicians is that the clinical algorithm and AI allowed doctors to reflect on or confirm their decision-making. One doctor gave an example of this, wherein they reviewed their decision based on the AI, “[I] looked at numbers and trends, [and the AI] gave me examples of why patients were better, worse, or the same”. Similarly, 2 physicians, 1 psychiatrist and 1 GP, reported that they would consider the AI recommendations if patients weren’t responding to current treatment; one psychiatrist stated “If the patient didn’t get better on current meds, then I would change the medication and look at what the algo said. Results were interesting, I haven’t used it a lot in practice but if my patient didn’t get well, then I would have use[d] the tool more”, though they also suggested the AI follow patients for a longer time (6 months) prior to making recommendations. Despite positive comments that the AI and CDSS provided “useful info, [and] remind[s] you treatment only has a chance of working. Maybe how long to try medication for before reassessing,” one psychiatrist also noted that when the difference in predicted remission between treatments was low that they were less likely to take this into account during decision making (as they put it, “on a population level, 5% [improvement in predicted remission] can make a big difference, but not for the person you’re treating that specific day”). Using the CDSS as a confirmation of clinical decisions was noted by 5/7 physicians (1 psychiatrist, 4 GPs). No physicians reported negative effects on patient outcomes as a result of using the tool.

One key aspect of assessing perceived clinical utility as well as barriers to use is considering whether or not users are willing to continue using the application in the future. 9 of 12 patients (6 patients of GPs; 3 patients of psychiatrists) reported that they would, as one patient put it, “like to continue using [the app] to monitor” how they are feeling. 2 patients reported they would use the app if there were changes to their treatment plan. 5 of 7 physicians said they were willing to continue using the tool after the study (2 GPs; 3 psychiatrists).

#### B. Impact on Physician-Patient Interaction and Relationship

All patients and clinicians noted neutral or positive impacts of the application on the physician-patient relationship. One psychiatrist shared that the app may have been beneficial in their doctor-patient relationship stating, “I felt like there was another outside kind of help and of objectification [which facilitated] the rapport with the patient”.

4 patients shared that the app in some way, as one patient put it, “enhanced” their relationship with their physician. One patient noted that the tool helped to facilitate conversations: “[the app] made it easier, as sometimes I have trouble verbalizing how I am feeling, so it helped the relationship.” The 3 other patients cited their increased involvement and understanding as the main factor for this change. One patient shared, “[using the app] made things a lot easier, […] not many of my other doctors have been able to make me feel good about treatment [and the app] definitely [had] a positive impact”. Similarly, a patient shared that “tracking symptoms/treatments helped [the] physician better manage patient care,” while another commented, “[the app] improved [the relationship by introducing] more knowledge and understanding in the relationship.” Other physicians and patients reported neutral impacts on the relationship.

There were comments supporting the use of the tool in facilitating shared decision making. For example, a clinician asserted that “it’s useful to have the feedback that the patient is improving or not. When I looked at the scales and saw that there was no improvement, I could speak about it with the patient and use that in my discussion with them”. 2 patients commented that they enjoyed the shared approach saying that they “appreciated that the doctor included [them] in app-use appointments” and that the app “improve[d] care on how they dealt with [their] depression together”.

##### Patient agency and self-management

Two physicians reported feeling that the app improves patient agency due to increased involvement required by questionnaires and the symptom tracking features of the app, with one physician stating, “[the app] improved things [by making] patient[s] more active participants [in their care and allowing them to] see objective changes on questionnaires”. Another physician expressed that the tool “made [the patient] more accountable” while another found the app to be “[of] benefit [as it] allowed patients to reflect on symptoms and see what type of questions others in their situation were being asked.” Mirroring this sentiment, the 9 patients who commented on patient agency all reported feeling that the app improved their agency and communication. For example, 1 patient credited the app with enabling him to be, “better able to self-reflect and articulate how he was feeling at visits,” a sentiment he shared with 3 additional patients. These patients communicated that the app gave them the opportunity to divulge certain information they might not have otherwise shared because they didn’t know it was relevant, because they found the experience difficult to express in words, or because they “don’t feel comfortable expressing [these experiences] out loud”.

#### C. Experience of Measurement-Based Care

Regularly responding to and reviewing standardized questionnaires, while potentially familiar as a concept, was a novel experience for many physicians and patients in the study, and constituted the most significant interaction with the tool in terms of time for patients, as well as an integral part of tool use for physicians.

##### Measurement-Based Care: Patient Perspective

The majority of patients (n=10) reported an increased awareness of their thoughts, feelings, and how they changed over time.

Patients generally found the questionnaires rapid and easy to complete; additionally, many reported enjoying or finding it helpful to take the time to reflect on how they were feeling (Box 1).

While 6 of 12 patients expressed, as one patient put it, that the “app covered everything [they] would have wanted to report,” a number of patients did find the use of standardized questionnaires to be restrictive. As will be discussed below, this is a reasonable tension to expect when using standardized questionnaires.

##### Measurement-Based Care: Physician Perspective

One physician said that the tool “made it somewhat easier to get standardized questionnaires completed by my patients [which is an] incentive [for use]”. Two other physicians agreed the questionnaires were beneficial in making patients more active in their care. Refer to *Patient agency and self-management* for specific quotes. Another said that it was “useful to monitor variations in scores”. Mirroring the concerns of some patients, one physician explained that many questionnaires are not always specific to the patient. In addition, there can be tension between the results of a standardized questionnaire and clinical judgement: one doctor described that in one instance “what the patient reported to me and their score on the tool did not add up. [Their] score was higher than the symptoms the patient was presenting. [On the other hand], the numbers produced by the AI/predictive model were fine”.

#### D. Understanding and Trust

##### Perceived level of Understanding of AI

Physicians had mixed responses when asked if they understood how the AI worked. 3/7 doctors (2 psychiatrists, 1 gp) stated they understood the AI to at least a basic level, while 3/7 doctors (1 psychiatrist, 2 gp) shared they felt they did not understand the AI. Generally, physicians reported using the AI as “a supplement to clinical judgement” and, as a result, they did not place emphasis on the importance of their understanding. A key point to note here is that, even if they did not understand the technical details in full, physicians understood the *role* of the AI as an assistant to their decision making. Refer to Supplementary Material for further comments.

##### Feelings of Trust in the CDSS

Physician trust in the CDSS, including the AI results, despite the mixed results regarding understanding of the AI component, was found to be generally positive. 6/7 physicians (2 psychiatrists, 4 GPs) stated that they trusted the CDSS; 1 psychiatrist with complex patients expressed hesitance in fully trusting the tool because it did not take into consideration failed treatments (as discussed above). In addition, as noted above in the section on clinical utility, the majority of physicians trusted the tool enough to use it to confirm their decisions, and 4/7 physicians reported being more open to prescribing medications they would not have considered otherwise.

Further, three doctors attributed their trust in the tool to the fact that the app mirrors the CANMAT guidelines and that the AI demonstrated “reasonable options that [they] would’ve suggested.” 5/7 doctors reported they trusted the tool because, as one doctor put it, it “was aligned with doctor’s clinical opinions and was good reinforcement from an exogenous source.” Doctors felt comfortable using the app in practice because they were, as per one physician, “never surprised” by the CDSS’ recommendations and the “AI explained reasoning behind its choices.” A physician noted that the CDSS reported recommendations that “felt accurate and informative.” Confirming the doctor’s initial thinking contributed to trust in the app and its continued use.

Patient feelings of how well the CDSS represented their individual clinical situation seemed to be an important component of their overall trust in the tool. 6/12 patients reported generally trusting the tool and feeling represented by it. One patient reported that “I found that it was pretty spot on and accurate with regards to how I felt. It almost seems like it had a good idea of who I am.” 6/12 patients felt reassured that the tool produced personalized results, with one patient sharing that it “felt personalized.” Another two patients shared that they felt that the tool is trustworthy and a “Good tool in the hands of a good doctor. I would be a little perturbed if a doctor based the diagnosis/medications solely on the app without going into depth with the patient.” Patients felt comfortable in the tool being used by physicians but “[they] don’t think the AI is capable of making a decision for the treatment of the patient.”. 6/12 patients stated that they trusted their doctors to “recognize when they should trust the AI.” Finally, one patient found it interesting to see their medication in the tool and to know that the choice of treatment was “backed by patient samples”.

## Discussion

In this paper, we present a mixed-methods feasibility study of novel AI-enabled CDSS. Seven physicians and 14 patients completed this feasibility study, conducted during the COVID-19 pandemic, and provided valuable insights regarding the feasibility, clinical utility and useability of the system; its impact on the physician-patient relationship; their experience of measurement-based care; and their understanding and trust in the CDSS. These different thematic categories, and relevant quantitative data, provide valuable insight into not only the integration of this particular CDSS into primary and specialty care, but into the general considerations for the integration of CDSS with AI components in mental healthcare.

### Clinical Feasibility, Utility and Useability

As noted in the quantitative results and the results of our previous paper on this study, which was focused on feasibility, (Popescu et al., 2021), both patients and physicians found the tool easy to use and feasible in a clinical setting, building on the results from our previous simulation-center based ease-of-use study (Benrimoh et al., 2021; Tanguay-Sela et al., 2021). Refer to supplementary material for further comments.

Physicians and patients agreed about the clinical utility of the application’s mood tracking features to provide improved information about patient status. Physicians appreciated the clinical algorithm and the AI/predictive model; 6/7 clinicians noted that it was useful in their clinical decision-making and a majority noted its utility to confirm clinical decisions. A number of physicians also noted its ability to help them reflect on clinical decisions or consider options they otherwise would not have, which may be an important contribution given the fact that physicians tend to use the same treatments repeatedly (Frank & Zeckhauser, 2007). Given that one key objective of this tool is to help personalize treatment, it is encouraging to recognize that physicians are open to considering the information it provides and re-visiting existing patterns (Henshall et al., 2017). It is also interesting to note that updated versions of the tool may have a role to play in providing continuing medical education and that this may in turn improve the utility of the clinical decision support. This is fundamentally how the system is intended to be used: rather than physicians abdicating responsibility and simply following AI recommendations, they are meant to incorporate the predictions of the model while still considering their own expertise and collaborating with the patient.

Other comments from physicians and patients also provide valuable insight into how to continue evolving AI-enabled CDSS’ to provide further clinical utility. For example, as noted above, one psychiatrist with complex patients noted that he would have preferred if the AI had taken previous treatment failure into account. While this was not possible with the current dataset and model architecture, it is something that will be taken into account during the design of future AI modules. The same psychiatrist noted that they would be more likely to use AI predictions if the predicted efficacy of treatments varied more greatly at the individual patient level. This bears further investigation, and this metric will also be used when selecting optimal models for inclusion in the CDSS in future releases. As noted above, some patients noted that they wanted to continue using the tool in a maintenance capacity, rather than with the same weekly intensity used during the study. The tool is indeed designed to allow for a reduction in the frequency of questionnaires, the questionnaire frequency was kept constant throughout the duration of the study in order to standardize assessments across patients. Further work on features supporting patients in maintenance might also extend the useful lifespan of the tool for each patient.

Physicians noted (see supplementary materials), in a manner similar to comments made in our simulation center study (Benrimoh et al., 2020), that they felt that more practice with the tool would have improved their comfort with it and ability to use it clinically. The protocol and application training session in this study for physicians was longer than in the simulation center study (roughly 1 hour compared to roughly 40 minutes); however, it appears that more practice would be beneficial. As a result, in our upcoming effectiveness study clinicians will receive more detailed training and will be encouraged by local coordinators to practice using the tool in the time between initial training and patient recruitment.

### Impact on Physician-Patient Relationship

The prospect of implementing a CDSS with a novel AI component raised concerns about potential deleterious effects on the clinician-patient relationship. Indeed, one reason that this study was designed with a baseline session without the tool followed by the introduction of the tool was to provide an opportunity to note if tool introduction caused friction between patients and physicians. Encouragingly, quantitative results from the end questionnaire and STAR questionnaires demonstrate no negative effects on the relationship, and this is echoed by the qualitative results.

While many clinicians and patients noted no change in the relationship-likely because, as noted by some patients above, the patients had existing positive relationships with their clinicians- there was a significant portion of patients (46%) who noted an improvement in their relationship with their physician. As such, the tool appears to either have no negative effect on the clinician-patient relationship, or to have a positive effect for nearly half of patients. The qualitative results provide two potential mechanisms by which this positive effect might be produced. One is improved communication (Lin, 2012), with patients feeling that their physicians had a better sense of their cases and were more prepared for appointments. The other is an improved sense of patient ability to self-advocate, which corresponded to physician comments about more active patient involvement in care, having the same questionnaire results and graphs available to both parties may have helped to improve this sense of patient self-advocacy (Walker et al., 2011). Both mechanisms may have contributed to the improvements in shared decision making commented on by participants.

This study was conducted during the COVID-19 pandemic; as a result, most visits were over the phone or using a teleconferencing solution. As detailed in the Supplementary Results, many participants noted that the tool supported the transition to telemedicine.

### Experience of Measurement-Based Care

The experience of measurement-based care was overall much appreciated by both physicians and patients, despite the need to answer four questionnaires weekly in addition to a number of baseline questionnaires. As noted, 6/7 physicians felt the tool helped them get a better sense of patient situations, and as discussed above patients felt their physicians were better prepared for their appointments. Furthermore, patients noted that an important aspect of using the tool for them was the chance to self-reflect and realize how their situations have evolved over time. They noted the utility of the CDSS as effectively a neutral party they could report symptoms to that were otherwise difficult to verbalize or discuss directly in sessions. These important benefits help to explain the positive impacts of the tool noted by participants on shared decision making and communication.

While the tool seems to help patients report and reflect on their clinical symptoms and course, its reliance on standardized questionnaires, necessary for effective measurement-based care (Fortney et al., 2017), does create some expected tensions. Not all patients felt the questionnaires allowed them to fully report how they were feeling, and a number of patients suggested improvements such as daily mood tracking or a free-form diary or comments section - as one patient put it, “something a little more personalized where you can write yourself”. Future versions of the tool may be able to incorporate more personalized elements, as well as to borrow tools from other methodologies, such as ecological momentary assessment, to begin personalizing patient data collection (Moskowitz & Young, 2006). This may in turn help support a wider range of therapies (i.e. patient diaries in cognitive behavioral therapy) (Nes et al., 2013).

### Understanding and Trust

Physician and patient understanding and trust of the app and AI were two significant factors queried in this study. Successful integration of the app and AI into standard practice is contingent on physicians’, as well as the patients’ perception of their physicians’ understanding and trust in the tool. Physicians and patients were not given explicit instructions on how to implement the tool and AI into their care, and physicians were not told how to talk to their patients about the CDSS. As such, we were able, as in our simulation center study, to study how physicians and patients naturally learned to use the tool in their interactions and how they included it in their understanding of the treatment process.

It is interesting to observe that neither physicians nor patients endorsed fully understanding the AI and how it worked, though a number of physicians did note a basic understanding (we note that all physicians were trained on the basics of how the AI model was trained and had the opportunity to ask questions during their training session). The patient’s reaction to their level of understanding was essentially to note that their main concern was their physician’s ability to understand and use the system appropriately. This underscores the importance of trust and strength in the relationship between physician and patient and, as such, makes it doubly important to demonstrate, as we have here, the lack of negative impact on the physician-patient relationship of the CDSS. Despite having only a basic understanding of the AI, the majority of physicians did endorse trusting the CDSS as a whole. Some clinicians noted that this was because the majority of information presented in the application (with the exception of predicted remission probabilities) came from established treatment guidelines, underscoring the value of the design of this CDSS, which sought to use AI to build upon the gold-standard approach. While they may not have understood all the *technical* details of the AI, clinicians *did* understand the way in which the AI was meant to be used- as an aid to clinical decision, as one more piece of information they could consider (Jeffries et al., 2021). Most physicians had only a few patients in this study. As such, one might argue that what was observed in this study is the ‘exploratory’ phase of physicians integrating this new technology - with physicians willing and able to use it, but still learning how it works and how best to leverage it in their practices. Given that technology remains quite novel to physicians, with a systematic review reporting that 70.2% of physicians had never used applications in clinical practice before (Kerst et al., 2020), it is important to provide continuing education and training for healthcare providers (Briganti & Le Moine, 2020). Our upcoming effectiveness trial, which asks physicians to enroll double the number of patients as in this study, may therefore provide more information about how physicians use this technology with more practice and practical experience with the system.

## Limitations

This study has a number of important limitations. One key limitation was the small sample size, which may limit the generalizability of results. In addition, participant interviews were recorded using written notes rather than voice recordings. The study was designed with a focus squarely on feasibility, with the tool being introduced only after the first visit with the patient. Indeed even patients noted this, with one patient mentioning how their medications had been changed prior to the introduction of the tool and that as such the CDSS likely did not have an impact at the level of treatment choice. We note that this was not intentional, as this study was not meant to test tool effectiveness but rather to determine if its introduction was feasible or if it had any deleterious effects on the clinician-patient relationship. This limitation is further discussed in Popescu et al., 2021. Finally, as noted above this study was adjusted in order to conduct it entirely virtually, due to the COVID-19 pandemic. One negative aspect of telemedicine was reduced opportunity for physicians to look at their screens together with patients, as many of the telemedicine follow-ups were conducted over the phone. During the simulation center study of this tool (Benrimoh et al., 2020), looking at screens together was an important part of the patient-physician interaction when using the tool. As such, this study may have benefitted from more in-person visits; on the other hand, the significantly positive experience of the tool supplementing telemedicine demonstrates both the versatility of the CDSS and the manner in which patients and clinicians can adapt its use to meet present circumstances.

## Conclusion

In conclusion, this paper has described a mixed-methods approach to the study of a novel AI-enabled CDSS for the treatment of major depression. Key themes regarding clinical utility, ease of use, feasibility, the impact on the physician-patient relationship, the experience of measurement-based care, and physician and patient understanding of and trust in the tool were discussed. Importantly, clinicians saw the tool as being a trustworthy support to help them explore more treatment options, and patients found the experience of measurement-based care to be one that helped improve self-reflection and support self-advocacy. A number of key learnings, such as the need for increased training time and the importance of administrative support, will help support future effectiveness trials of this - and potentially, of other - clinical decision support systems in mental healthcare and beyond. Next steps also include upgrades to the AI model, such as an AI that takes individual failed treatment history and patient trajectories into account.

## Data Availability

All data produced in the present study are available upon reasonable request to the authors

## Acknowledgements

We thank patients, physicians, and local staff for participating in this feasibility study.

## Notes

### Competing Interest Statement

D.B., C.A., R.F., S.I., and K.P. are shareholders and either employees, directors, or founders of Aifred Health. J.M. is employed by Aifred Health. M.T.S. is employed by Aifred Health and is an options holder. S.Q., G.G., D.S., M.S., K.W., T.P., E.S., E.L., MJ. F., J.B., B.D., and C.P., have been or are employed or financially compensated by Aifred Health. S.P., K.H., J.K. are members of Aifred Health's scientific advisory board and have received payments or options. W.S. is a member of the data safety monitoring board. H.M. has received honoraria, sponsorship or grants for participation in speaker bureaus, consultation, advisory board meetings and clinical research from Acadia, Amgen, HLS Therapeutics, Janssen-Ortho, Mylan, Otsuka-Lundbeck, Perdue, Pfizer, Shire and SyneuRx International. S.R reports owning shares in Aifred Health. All other authors report no relevant conflicts.

### Clinical Trial

NCT04061642

### Funding Statement

Funding was obtained between initial IRB approval (July 29th, 2019) and the start of the study but was disclosed to REB as amendments were approved prior to study start.
Funding sources: Aifred Health Inc.; Innovation Research Assistance Program, National Research Council Canada; ERA-Permed Vision 2020 grant supporting IMADAPT; Government of Quebec Nova Science; MEDTEQ COVID-Relief Grant.

### Author Declarations

This research was reviewed and approved by the Research Ethics Board of the Douglas Mental Health University Institute, Montreal West Island Integrated University Health and Social Services Centre.

## References

Adli, M., Wiethoff, K., Baghai, T. C., Fisher, R., Seemüller, F., Laakmann, G., Brieger, P., Cordes, J., Malevani, J., Laux, G., Hauth, I., Möller, H. J., Kronmüller, K. T., Smolka, M. N., Schlattmann, P., Berger, M., Ricken, R., Stamm, T. J., Heinz, A., & Bauer, M., 2017. How Effective Is Algorithm-Guided Treatment for Depressed Inpatients? Results from the Randomized Controlled Multicenter German Algorithm Project 3 Trial. The international journal of neuropsychopharmacology. 20(9), 721–730. https://doi.org/10.1093/ijnp/pyx043

American Psychiatric Association. DSM-5 Diagnostic Classification. Diagnostic and Statistical Manual of Mental Disorders. 10th ed.; 2013.

Asan, O., Bayrak, A. E., & Choudhury, A. 2020. Artificial Intelligence and Human Trust in Healthcare: Focus on Clinicians. Journal of medical Internet research, 22(6), e15154. https://doi.org/10.2196/15154

Bate, L., Hutchinson, A., Underhill, J., & Maskrey, N., 2012. How clinical decisions are made. British journal of clinical pharmacology. 74(4): 614–620. https://doi.org/10.1111/j.1365-2125.2012.04366.x

Bauer, M., Pfennig, A., Linden, M., Smolka, M. N., Neu, P., & Adli, M. 2009. Efficacy of an algorithm-guided treatment compared with treatment as usual: a randomized, controlled study of inpatients with depression. Journal of clinical psychopharmacology. 29(4): 327–333

Benrimoh D, Fratila R, Israel S, Perlman K, Mirchi N, Desai S, Rosenfeld A, Knappe S, Behrmann J, Rollins C, You RP. Aifred health, a deep learning powered clinical decision support system for mental health. InThe NIPS’17 Competition: Building Intelligent Systems 2018 (pp. 251–287). Springer, Cham

Benrimoh D, Tanguay-Sela M, Perlman K, Israel S, Mehltretter J, Armstrong C, Fratila R, Parikh SV, Karp JF, Heller K, Vahia IV. Using a Simulation Centre to Evaluate the Effect of an Artificial Intelligence-Powered Clinical Decision Support System for Depression Treatment on the Physician-Patient Interaction. medRxiv. 2020 Jan 1.

Blumenthal, S.R., Castro, V.M., Clements, C.C., Rosenfield, H.R., Murphy, S.N., Fava, M., Weilburg, J.B., Erb, J.L., Churchill, S.E., Kohane, I.S. and Smoller, J.W., 2014. An electronic health records study of long-term weight gain following antidepressant use. JAMA psychiatry. 71(8), 889–896. doi:10.1001/jamapsychiatry.2014.414

Bohn, M. J., Babor, T. F., Kranzler, H. R., 1995. The Alcohol Use Disorders Identification Test (AUDIT): validation of a screening instrument for use in medical settings. J Stud Alcohol. 56(4), 423–432. doi:10.15288/jsa.1995.56.423

Braun, V. & Clarke, V., 2006. Using thematic analysis in psychology. Qualitative Research in Psychology. 3(2)77–101. DOI: 10.1191/1478088706qp063oa

Brigant, G. & Le Moine O., 2020. Artificial intelligence in medicine: today and tomorrow. Frontiers in medicine. 7, 27. https://doi.org/10.3389/fmed.2020.00027

Bussone A, Stumpf S, O’Sullivan D. The role of explanations on trust and reliance in clinical decision support systems. In2015 international conference on healthcare informatics 2015 Oct 21 (pp. 160–169). IEEE

Canadian Institutes of Health Research, Natural Sciences and Engineering Research Council of Canada, and Social Sciences and Humanities Research Council (2005), Tri-Council Policy Statement: Ethical Conduct for Research Involving Humans.

Fortney, J. C., Unützer, J., Wrenn, G., Pyne, J. M., Smith, G. R., Schoenbaum, M., & Harbin, H. T. A Tipping Point for Measurement-Based Care. Psychiatric services (Washington, D.C.) 2017; 68(2): 179–188. https://doi.org/10.1176/appi.ps.201500439

Frank, R. G., & Zeckhauser, R. J., 2007. Custom-made versus ready-to-wear treatments: behavioral propensities in physicians’ choices. Journal of health economics. 26(6), 1101–1127. https://doi.org/10.1016/j.jhealeco.2007.08.002

Gille F, Jobin A, Ienca M., 2020. What we talk about when we talk about trust: Theory of trust for AI in healthcare. Intelligence-Based Medicine. 1, 100001.

Goldberg, S. B., Buck, B., Raphaely, S., & Fortney, J. C., 2018. Measuring Psychiatric Symptoms Remotely: a Systematic Review of Remote Measurement-Based Care. Current psychiatry reports. 20(10), 81. https://doi.org/10.1007/s11920-018-0958-z

Henshall C, Marzano L, Smith K, Attenburrow MJ, Puntis S, Zlodre J, Kelly K, Broome, MR, Shaw S, Barrera A, Molodynski A. 2017. A web-based clinical decision tool to support treatment decision-making in psychiatry: a pilot focus group study with clinicians, patients and carers. BMC psychiatry. 17(1), 1–0.

Jeffries M, Salema NE, Laing L, Shamsuddin A, Sheikh A, Avery A, Chuter A, Waring J, Keers RN., 2021. The implementation, use and sustainability of a clinical decision support system for medication optimisation in primary care: A qualitative evaluation. PloS one. 16(5), e0250946.

Jones, C., Thornton, J., & Wyatt, J. C., 2021. Enhancing trust in clinical decision support systems: a framework for developers. BMJ health & care informatics. 28(1), e100247. https://doi.org/10.1136/bmjhci-2020-100247

Juravle, G., Boudouraki, A., Terziyska, M., & Rezlescu, C., 2020. Trust in artificial intelligence for medical diagnoses. Real-World Applications in Cognitive Neuroscience. 253, 263–282.

Katz, A. J., Dusetzina, S. B., Farley, J. F., Ellis, A. R., Gaynes, B. N., Castillo, W. C., Stürmer, T., Hansen, R. A., 2012. Distressing adverse events after antidepressant switch in the Sequenced Treatment Alternatives to Relieve Depression (STAR*D) trial: influence of adverse events during initial treatment with citalopram on development of subsequent adverse events with an alternative antidepressant. Pharmacotherapy. 32, 234–43.

Kennedy, S. H., Lam, R. W., McIntyre, R. S., Tourjman, S. V., Bhat, V., Blier, P., Hasnain, R. M., Jollant, F., Levitt, A. J., MacQueen, G. M., McInerney, S. J., McIntosh, D., Milev, R. V., Müller, D. J., Parikh, S. V., Pearson, N. L., Ravindran, A. V., Uher, & CANMAT Depression Work Group., 2016. Canadian Network for Mood and Anxiety Treatments (CANMAT) 2016 Clinical Guidelines for the Management of Adults with Major Depressive Disorder: Section 3. Pharmacological Treatments. Canadian journal of psychiatry. Revue canadienne de psychiatrie. 61(9), 540–560. https://doi.org/10.1177/0706743716659417

Kerst, A., Zielasek, J., Gaebel, W., 2020. Smartphone applications for depression: a systematic literature review and a survey of health care professionals’ attitudes towards their use in clinical practice. Eur Arch Psychiatry Clin Neurosci. 270(2), 139–152. doi:10.1007/s00406-018-0974-3

Kroenke, K., Spitzer, R. L., Williams, J. B. W., 2001. The PHQ-9: Validity of a brief depression severity measure. J Gen Intern Med. 16(9), 606–613. doi:10.1046/j.1525-1497.2001.016009606.x

Lin, H. C. 2012. Real-time clinical decision support system. Medical Informatics. 111–36.

Scale McGuire-Snieckus, R., McCabe, R., Catty, J., Hansson, L., Priebe, S., 2007. A new to assess the therapeutic relationship in community mental health care: STAR. PsycholMed. 37(1), 85–95. https://doi.org/10.1017/s0033291706009299

Merlhiot, G., 2014. Adaptation and Validation of the Standardized Assessment of Personality – Abbreviated Scale as a Self-Administered Screening Test (SA-SAPAS). J Psychol Psychother. 04(06). doi:10.4172/2161-0487.1000164

Moskowitz, D. S., & Young, S. N. (2006). Ecological momentary assessment: what it is and why it is a method of the future in clinical psychopharmacology. Journal of psychiatry & neuroscience : JPN. 31(1), 13–20.

Nes, A. A., Eide, H., Kristjánsdóttir, Ó. B., & van Dulmen, S., 2013. Web-based, self-management enhancing interventions with e-diaries and personalized feedback for persons with chronic illness: a tale of three studies. Patient education and counseling. 93(3), 451–458. https://doi.org/10.1016/j.pec.2013.01.022

Patel, S. R., Bakken, S., & Ruland, C., 2008. Recent advances in shared decision making for mental health. Current opinion in psychiatry. 21(6), 606–612. https://doi.org/10.1097/YCO.0b013e32830eb6b4

Popescu, C., Golden, G., Benrimoh, D., et al. (2021). Evaluating the Clinical Feasibility of an Artificial Intelligence-Powered, Web-Based Clinical Decision Support System for the Treatment of Depression in Adults: Longitudinal Feasibility Study. JMIR formative research, 5(10), e31862. https://doi.org/10.2196/31862

Scott, K., & Lewis, C. C., 2015. Using Measurement-Based Care to Enhance Any Treatment. Cognitive and behavioral practice. 22(1), 49–59. https://doi.org/10.1016/j.cbpra.2014.01.010

Sim, I., Gorman, P., Greenes, R. A., Haynes, R. B., Kaplan, B., Lehmann, H., & Tang, P. C., 2001. Clinical decision support systems for the practice of evidence-based medicine. Journal of the American Medical Informatics Association : JAMIA. 8(6), 527–534. https://doi.org/10.1136/jamia.2001.0080527

Skinner, H. A., 1982. The drug abuse screening test. Addict Behav. 7(4), 363–371. doi:10.1016/0306-4603(82)90005-3

Spitzer, R. L, Kroenke, K., Williams J. B. W., Löwe B., 2006. A Brief Measure for Assessing Generalized Anxiety Disorder: The GAD-7. Arch Intern Med. 166(10), 1092. doi:10.1001/archinte.166.10.1092

Sutton, R.T., Pincock, D., Baumgart, D.C. et al., 2020. An overview of clinical decision support systems: benefits, risks, and strategies for success. npj Digit. Med. 3, 17. https://doi.org/10.1038/s41746-020-0221-y

Tanguay-Sela, M., Benrimoh, D., Popescu, C., Perez, T., Rollins, C., Snook, E., … Margolese, H. (2021, January 25). Evaluating the Perceived Utility of an Artificial Intelligence-Powered Clinical Decision Support System for Depression Treatment Using a Simulation Centre. Retrieved from https://www.medrxiv.org/content/10.1101/2021.04.21.21255899v1.full-text

Tran, B. X., McIntyre, R. S., Latkin, C. A., Phan, H. T., Vu, G. T., Nguyen, H., Gwee, K. K., Ho, C., & Ho, R., 2019. The Current Research Landscape on the Artificial Intelligence Application in the Management of Depressive Disorders: A Bibliometric Analysis. International journal of environmental research and public health. 16(12), 2150. https://doi.org/10.3390/ijerph16122150

Walker, J., Leveille, S. G., Ngo, L., Vodicka, E., Darer, J. D., Dhanireddy, S., Elmore, J. G., Feldman, H. J., Lichtenfeld, M. J., Oster, N., Ralston, J. D., Ross, S. E., & Delbanco, T., 2011. Inviting patients to read their doctors’ notes: patients and doctors look ahead: patient and physician surveys. Annals of internal medicine. 155(12), 811–819. https://doi.org/10.7326/0003-4819-155-12-201112200-00003

Wisniewski, S. R., Rush, A.J., Balasubramani, G. K., Trivedi, M. H., Nierenberg, A. A., 2006. Self-Rated Global Measure of the Frequency, Intensity, and Burden of Side Effects: J Psychiatr Pract.12(2), 71–79. doi:10.1097/00131746-200603000-00002

Yoshino, A., Sawamura, T., Kobayashi, N., Kurauchi, S., Matsumoto, A., & Nomura, S., 2009. Algorithm-guided treatment versus treatment as usual for major depression. Psychiatry and clinical neurosciences. 63(5), 652–657. https://doi.org/10.1111/j.1440-1819.2009.02009.x

