## Supplementary Material for "A Mixed-Methods Feasibility Study of a Novel AI-Enabled, Web-Based, Clinical Decision Support System for the Treatment of Major Depression in Adults"

QUALITATIVE ANALYSIS

Data from 7 physicians and 12 patient end interviews was collected and analyzed to assess perceived clinical utility, trust and understanding, and perceived impact on patient-physician relationship of the clinical decision support system (CDSS).

The physician themes consisted of 16 sub-themes, the patient category had 15 sub-themes, both the physician-patient relationship (interaction) and COVID-19 categories had 5 sub-themes and the miscellaneous category had 2 sub-themes.

Physician sub-themes: Subjective view on apt. Length, Ease of use (general), Impact of time spent with app on use of app/comfort with app, Incentive to use tool & motivation, Willingness to use the tool after study - burdensome? Tedious?, Understanding of AI, (Physician's perception of) patients understanding of AI, Physician perception of clinical utility for patients (how physician felt tool affected patient), Clinical effectiveness - openness to prescribe new meds based on AI, Clinical effectiveness - other factors influencing whether the physician used the tool or not (general), Physician use of tool with different patients (eg. of different severities), Trust in AI, Level of agreement of predicted medication via algo, Feeling that clinical/treatment decisions were confirmed (or not) by tool’s suggestions/felt like the tool supplemented clinical knowledge, Frequency of app use, Questionnaire use/comments on questionnaires, App improvement suggestions

Patient sub-themes: Subjective view on appointment length, Ease of use, Motivation to use tool/enjoyment of tool use, Frequency of use, time spent using app, Interactions with the tool - interest in using it, use of reminders, convenience, frustrations, Interactions with the tool - comments about info asked + presented, Tool’s impact on symptomatology and introspection/awareness of symptoms	Interest/willingness in continuing app use post study, Introspection regarding their diagnosis/treatment, General comments regarding treatment (changes, side effects, etc.), Interest in study/contribution to science, Clinical effectiveness - impact of tool on care, Understanding of AI, (Patient’s perception of) Physician understanding of AI/tool, Trust in the tool, Privacy concerns regarding tool, App improvement suggestions

Interaction sub themes: Impact of app on relationship in general, In-visit app use, Patient agency: Self-management and self-advocacy (better communicate how they’re feeling, be more active in their care?), Did the tool influence how the physician conducted their appointments (IE. What questions were asked, whether deeper conversations were had RE. Symptoms, was more information shared by patient, etc.), Comments related to shared decision making

COVID/ Telemedicine sub themes: Impact of COVID-19 on care, Impact of COVID-19 on individual, Impact of tool on care in the context of COVID-19, Tool used for telemedicine, Evolution of tool use throughout COVID (transition from in-person to telemedicine)

Miscellaneous sub themes: Mention of psychotherapy (wanting more data), RA reminders

A. CLINICAL UTILITY AND USEABILITY OF THE APP

*Ease of Use*

Overall subjects noted the tool was easy to use. 10 of 12 patients reported that the app was easy to use. No patients discussed difficulties using the technology. 6 of 7 physicians (4 GPs; 2 psychiatrists) reported the tool was easy to use, with one general practitioner detailing the tool was “easy and user-friendly, steps and suggestions are well explained.” 2 of these physicians (2 psychiatrists) also reported that using the tool felt like a chore, with one specifying that this was due more to patients not completing the required questionnaires than to the tool itself. 3 physicians (1 GP; 2 psychiatrists) reported they would have felt more comfortable with the tool with more time and practice. “There’s room for improvement with more practice,” one physician (psychiatrist) commented, “I would have liked to practice more” prior to and in proximity to their first patient entering the study. Interestingly, 2 of these 3 physicians also asserted that they think more practice and increased comfort with the tool could lead to better care, thus justifying the extra cost of time for training.

*Barriers to Use*

Potential barriers to use identified included privacy concerns, ease of account creation, and integration into EMR systems. Two physicians cited challenges specifically with respect to account setup and linking (1 GP; 1 psychiatrist), and 2 physicians (2 GP) suggested that the tool be integrated with EMR to streamline use. These barriers will be discussed further below.

No patients had subjective privacy concerns, but 4 patients speculated that other patients may not be comfortable providing personal information, which could represent a barrier to use. For instance, one patient noted “I’m not as security-conscious as some people I know. I think it really depends on the type of person.”

1 physician (GP) said they would not continue to use the tool because of the burden, described above, involved in linking patient and doctor accounts. The seventh physician did not comment specifically on whether they would continue using the tool after the study.

*Clinical Utility*

With respect to the questionnaires responded to by patients in the tool, 5 physicians (3 GP; 2 psychiatrists) reported that they were helpful and/or added value. 1 physician (GP) credited the tool with providing questionnaires they may not have thought to use otherwise. 1 physician (psychiatrist) said that they used the basic questionnaires (the GAD-7 and PHQ-9) but that the range of other questionnaires beyond that were “too cumbersome.” The way the questionnaires impacted clinical decision-making varied among physicians. For example, despite the fact that “reading the change in scores [prior to appointments] definitely gave some orientation as to where the patient is going and if things are improving or not”, one doctor (psychiatrist) did “not think it would have changed my decision making during the encounter”. This physician also noted, however, that it was “useful to have the feedback that the patient is improving or not.” On the other hand, one physician (GP) commented on the value of the tool to “objectify scores” and said it “allowed [the] physician to think more about clinical decision-making.”

The clinical algorithm noted that psychotherapy could be effective for some patients, which was met with mixed reactions. One patient had previously tried therapy and found it ineffective, while two patients were pleased with the suggestion, with one expressing that “he was hesitant to go back on medication so he liked that the app did not suggest pharmacotherapy. He was able to get a therapist out of the experience and that has worked really well for him.”

*Clinical Utility – Telemedicine*

This study coincided with the onset of the COVID-19 pandemic in Canada and the subsequent lockdowns. As such the interview was amended in order to add questions about the utility of the tool in the context of telemedicine. Generally, physicians and patients found the tool to be helpful in the context of the transition to telemedicine, noting that it facilitated patient follow-up, helped reduce the impersonal aspect of telemedicine, and created the sense that both patients and physicians were more prepared for visits. 9 of 12 patients (5 patients of GPs; 4 patients of psychiatrists) and 4 of 7 physicians (3 GPs; 1 psychiatrist) reported positive experiences with the tool in the context of telemedicine. One physician (GP) noted that the “Tool was perfect for use in COVID!” while another physician (GP) commented, “Telemedicine was facilitated with tool, such a good idea for [an] app.” The latter also reported the app’s value in saving time for telehealth.

A theme emerged amongst both physicians and patients of the role of questionnaires in making patients feel that both they and their physician were more prepared for visits, and in giving physicians the ability to more effectively share patients’ progress with them. 3 physicians also commented that the tool helped facilitate the transition to telemedicine. One physician (GP) stated, “Think there is lots of room for improvement in telemedicine, but this tool was very helpful and a really good place to start!” Another of these physicians (GP) observed that the app “made the transition to telemedicine for treatment of mental health easier.” Another physician (psychiatrist) reported a similar perception of the app’s role saying, “The tool was complementary to the process of going online.” One patient observed that the “Tool facilitated care in telemedicine” and another stated that “The app definitely helped in the context that everything was done over telemedicine.”

2 patients commented on the tool’s value in increasing comfort with the otherwise impersonal telemedicine experience. One (with psychiatrist) stated, “I felt more comfortable sharing experiences over teleconference than I otherwise would have.” The other (with psychiatrist) observed, “Because everything was over the phone, [it was] a little bit impersonal. The tool helped in that regard.” One patient commented that the tool “Facilitated them [appointments] because he went into phone calls knowing what to say, as questionnaires let him think about it before.” Another patient commented on the physician’s preparedness for appointments, saying they felt “the tool facilitated the sessions as doctor could be more aware of how they were feeling between appointments.” 2 patients commented specifically on mood tracking with one saying that the tool “was helpful. Moves things along and made them aware of their moods since last call” and the other observed that the mood tracking of app was valuable especially since they cannot meet in person. One physician (psychiatrist), meanwhile, commented particularly on using the tool to share the results of this mood-tracking via questionnaires with their patients. They commented, “Tool used for almost all video follow-ups; was especially helpful if the patient did the rating scales. I was using screen share to show patients their scales and graphs which was quite useful.” 2 patients reported that the app helped to supplement aspects of their care that telemedicine detracted from in comparison with an in-person experience. One patient (with psychiatrist) reported, “Yes [COVID] had an effect in decreasing physician visits which I would have preferred. The app helped to palliate some of what was missing.” The other (with GP) observed, “I think it facilitated them because he was able to see those data points and comment on them. So it’s a way to substitute for seeing the patient [in-person] and seeing what they say vs. how they look; that is one set of data vs. this kind of data.” 1 patient (with psychiatrist) reported no difference in their experience of virtual versus in-person appointments with the tool.

The one physician (GP) who commented specifically on the tool’s role in in-person versus telemedicine interestingly evaluated the app as being perhaps better suited to telemedicine. “[It was] less awkward to do while on telemedicine, whereas in office might be a bit awkward” as they don’t like to be looking at a screen while in-person with a patient. On the other hand, one physician (GP) noted that, due to the challenge of the pandemic in that “any extra time needed for anything was a magnified burden.”

Given that many visits were conducted over the phone due to COVID-19, it was at times difficult for patients to know when their physician was consulting the algorithm or the tool in general. One patient expressed that they were “unsure if physician took tool into account in clinical decision making” and another “wasn’t made aware of the results [of the clinical algorithm]. He thinks it probably could have had an impact and maybe help[ed] [his physician] decide to switch antidepressants.”

B. IMPACT ON PHYSICIAN-PATIENT INTERACTION AND RELATIONSHIP

Another doctor described no impact on their current patient relationships but speculated that, “maybe if [I had had a] new patient it would [have impacted the relationship]”.

Similarly, a patient reflected:

"I think I had a different relationship with my GP than most people do so it did not change it that much because we have a very good and honest relationship. And I see a psychologist that he works with very closely so it wasn’t a huge impact because that relationship was already there. But it would be very useful for a new patient-physician relationship or a doctor without much time to give them info on when they’re not doing well and to take them seriously."

*Changes in the patient-clinician interaction*

Physicians had varied approaches to using the app during appointments with their patients. Eleven patients reported that their doctors referenced or shared results using the tool during their appointments, mostly verbally, with 7 patients commenting that their physicians explained what they were seeing on their screen during telehealth appointments, and one patient whose physician showed them their screen while using the tool in person.

The tool not only influenced how information was shared between patients and physicians, but the structure and content of appointments. Physicians expressed that the tool helped them conduct their appointments as the already-completed questionnaires saved time. Physicians also noted the utility of the CDSS in keeping them informed about how patients evolved over time.

One physician explained that they “would read back scores and mention what [the] app recommends” and another stated that she “explained what she was doing and seeing” during telehealth appointments, and that it was “nice to share thinking” during telehealth appointments. One physician found it “quite useful” to screen share to “show patients their scales and graphs”.

One physician felt that the information the application provided on the evolution of the patients’ symptoms allowed them to inform their discussions with patients and orient them to the direction the patient was headed in prior to appointments, stating that “reading the change in scores definitely gave some orientation as to where the patient is going and if things are improving or not.” Another physician echoed this sentiment, and added that the tool allowed them to think more about clinical decision making and work in “partnership with [the] patient” and confirm trends in symptoms, using the “tool without taking attention off of [the] patient.”

5 patients felt that their doctor was more aware of their current state of mind during appointments. In fact, one patient stated it allowed the doctor to “more easily understand how [they were] feeling” and two patients noted feeling that the doctor was more aware of their symptoms and situation prior to their visit.

C. EXPERIENCE OF MEASURMENT-BASED CARE

*Measurement-Based Care: Patient Perspective*

Three of 12 patients found that the questionnaires provided them perspective and the opportunity to reflect on how they were feeling during the study compared to other points in their lives. When looking back, one patient shared that “I sometimes felt that the depression wasn’t as bad as it had been before the study so I felt like ‘oh, I would’ve been a better candidate about a year ago when my depression was worse’.”

With respect to the adequacy of the questionnaires themselves, 6 of 12 patients (4 with GP; 2 with psychiatrist) expressed that the “app covered everything [they] would have wanted to report,” with one patient noting that they were “able to select the majority of [their] symptoms on the app so it was good for that,” and that they have many symptoms so this was important to them. On the other hand, 5 of 12 patients expressed that they were dissatisfied with the information they were able to report in the app, or felt that some of the questions did not pertain to them. Due to the use of standardized questionnaires, some patients felt that they were restricted in their ability to fully express how they were feeling. One patient noted that they “allowed her to report [the] majority of things most important to her but wished there were more answer options or comments at times,” and another “felt [that] some questions had an in-between response. Some questions [were] not applicable to her, but [she] had to fill out all questions to submit” the questionnaire. 3 of 12 patients (1 with GP; 2 with psychiatrist) found it difficult to answer questionnaires (e.g., PHQ-9) when they asked for reflection over the “last 2 weeks since it was too much time” to be able to provide accurate and precise responses.

D. UNDERSTANDING AND TRUST

*Perceived level of Understanding of AI*

One physician stated that “It matters a bit that I understand it, but then again, the way you can make your decisions is multifunctional”, implying that they considered the AI but also other aspects when making decisions (as was intended). Patient perception of physician understanding was varied. 7/12 patients felt that the physician understood the tool well enough to use in session. 4/14 patients (patients of GPs) were unsure whether their physicians understood the AI. Interestingly, no patients reported that their doctor did not understand the AI. 6/12 patients (2 patients of a psychiatrist, 4 patients of a GP) reported that it was more important that the physician understands the tool they are using more than for the patient to understand the AI. Patients generally reported mixed perceptions of their own understanding of the AI.

Patients also reported mixed understanding. 4/12 patients reported they had either a good idea or basic understanding of how the AI works. 6/12 patients shared that they had little understanding of the AI, however they were not particularly concerned, with one patient stating “As long as it works, it works – whether it’s magic or AI”. 2/12 patients (2 gp) reported they were unsure of their level of understanding of the AI, and one of those patients shared they did not put much thought into the AI. A common theme amongst patients was that they felt it was more important that their physician understood the AI than that they did. 7 physicians shared comments regarding their perception of patient understanding of the AI. 2/7 physicians (2 gp) shared that they believed their patients did not understand the AI. 3 physicians (2 psychiatrist, 2 gp) shared that their patients either did not care to understand the AI (1/12) or did not think about the AI (2/12). 2 doctors (1 psychiatrist, 1 gp) reported that they were unsure.

*Feelings of Trust in the CDSS*

Physicians were not required to use the app or apply its recommendations, so this comment shows that some doctors may have only trusted the tool if it agreed with them; this is perhaps reasonable caution in the context of a new technology, and may also represent reactions of physicians to an exogenous source of clinical input.

Interestingly, one doctor shared using the app as a test; “If [the] app agreed with me, I got the answer. If it disagreed, it was wrong.”

One patient noted concerns that it might be “biasing to read about symptoms on a questionnaire”, indicating, effectively, concerns about a nocebo […] effect of questionnaire responding. However, patients felt reassured that the AI produced personalized results, with one sharing that she “feels it represented her well.”

Discussion

*Clinical Feasibility, Utility and Useability*

Two key barriers to use were identified by physicians, however, which can help instruct future CDSS deployments. The first involved linking physician and patient accounts. The process for linking accounts in the study involved the patient or clinician generating a one-time code which was then shared directly with the other party to link the accounts. While secure and not dependent on local administrative support or IT infrastructure, this was, as one GP noted, “tedious”. As a result, an updated version of the tool which will be implemented in a future clinical trial has an administrator portal which requires the involvement of more personnel but eliminates this complexity as the administrator can link the accounts.

Physicians also suggested that the tool be integrated with the electronic medical record to streamline use; this was not possible during the study because of the discordance between IT infrastructure at the different sites in this study, but is planned for future implementations.

LEC-5 Questionnaire Results:

| Question Text | Patients with a positive response (%) n=13 |
| --- | --- |
| Natural disaster (for example, flood, hurricane, tornado, earthquake) | 2 (15.38%) |
| Fire or explosion | 5 (38.46%) |
| Transportation accident (for example, car accident, boat accident, train wreck, plane crash) | 9 (69.23%) |
| Serious accident at work, home, or during recreational activity | 4 (30.77%) |
| Exposure to toxic substance (for example, dangerous chemicals, radiation) | 2 (15.38%) |
| Physical assault (for example, being attacked, hit, slapped, kicked, beaten up) | 4 (30.77%) |
| Assault with a weapon (for example, being shot, stabbed, threatened with a knife, gun, bomb) | 2 (15.38%) |
| Sexual assault (rape, attempted rape, made to perform any type of sexual act through force or threat of harm) | 3 (23.08%) |
| Other unwanted or uncomfortable sexual experience | 9 (69.23%) |
| Combat or exposure to a war-zone (in the military or as a civilian) | 2 (15.38%) |
| Captivity (for example, being kidnapped, abducted, held hostage, prisoner of war) | 0 (0%) |
| Life-threatening illness or injury | 6 (46.15%) |
| Severe human suffering | 2 (15.38%) |
| Sudden violent death (for example, homicide, suicide) | 3 (23.08%) |
| Sudden accidental death | 7 (53.85%) |
| Serious injury, harm, or death you caused to someone else | 2 (15.38%) |
| Any other very stressful event or experience | 4 (30.77%) |

The LEC-5 is 17 questions long and asks patients about traumatic events. Answer options are the following: “Happened to me; Witnessed it; Learned about it; Part of my job; Not sure; Doesn’t apply”. Any response that was not “Doesn’t apply” or “Not sure” was counted as a positive response and is represented as a percentage in the table above.
